# Outcomes with and without outpatient SARS-CoV-2 treatment for patients with COVID-19 and systemic autoimmune rheumatic diseases: A retrospective cohort study

**DOI:** 10.1101/2022.10.27.22281629

**Authors:** Grace Qian, Xiaosong Wang, Naomi J. Patel, Yumeko Kawano, Xiaoqing Fu, Claire E. Cook, Kathleen M.M. Vanni, Emily N. Kowalski, Emily P. Banasiak, Katarina J. Bade, Shruthi Srivatsan, Zachary K. Williams, Derrick J. Todd, Michael E. Weinblatt, Zachary S. Wallace, Jeffrey A. Sparks

## Abstract

**Objective:** To investigate temporal trends, severe outcomes, and rebound among systemic autoimmune rheumatic disease (SARD) patients according to outpatient SARS-CoV-2 treatment.

**Methods:** We performed a retrospective cohort study investigating outpatient SARS-CoV-2 treatments among SARD patients at Mass General Brigham (23/Jan/2022-30/May/2022). We identified SARS-CoV-2 infection by positive PCR or antigen test (index date=first positive test) and SARDs using diagnosis codes and immunomodulator prescription. Outpatient treatments were confirmed by medical record review. The primary outcome was hospitalization or death within 30 days following the index date. COVID-19 rebound was defined as documentation of negative then newly-positive SARS-CoV-2 tests. The association of any vs. no outpatient treatment with hospitalization/death was assessed using multivariable logistic regression.

**Results:** We analyzed 704 SARD patients with COVID-19 (mean age 58.4 years, 76% female, 49% with rheumatoid arthritis). Treatment as outpatient increased over calendar time (p<0.001). A total of 426(61%) received outpatient treatment: 307(44%) with nirmatrelvir/ritonavir, 105(15%) with monoclonal antibodies, 5(0.7%) with molnupiravir, 3(0.4%) with outpatient remdesivir, and 6(0.9%) with combinations. There were 9/426 (2.1%) hospitalizations/deaths among those treated as outpatient compared to 49/278 (17.6%) among those with no outpatient treatment (adjusted odds ratio [aOR] 0.12, 0.05 to 0.25). 25/318 (8%) of patients who received oral outpatient treatment had documented COVID-19 rebound.

**Conclusion:** Outpatient treatment was strongly associated with lower odds of severe COVID-19 compared to no outpatient treatment. At least 8% of SARD patients experienced COVID-19 rebound. These findings highlight the importance of outpatient COVID-19 treatment for SARD patients and the need for further research on rebound.

**KEY MESSAGES:** 

**What is already known on this topic?:**

- Previous studies suggest that monoclonal antibodies are an effective outpatient treatment option for patients at high-risk of severe COVID-19, including those with systemic autoimmune rheumatic diseases (SARDs).
- Nirmatrelvir/ritonavir and molnupiravir are recently-authorized effective oral outpatient SARS-CoV-2 treatment options, but clinical trials were performed among the general population, mostly among unvaccinated and prior to Omicron viral variants.
- Oral outpatient SARS-CoV-2 treatments may result in COVID-19 rebound, characterized by newly-positive COVID-19 testing and recurrent symptoms, but no studies have investigated rebound prevalence among SARD patients.

**What this study adds?:**

- This is one of the first studies investigating outpatient SARS-CoV-2 treatments among SARD patients that includes oral options and quantifies the prevalence of COVID-19 rebound.
- Outpatient treatment was associated with 88% reduced odds of severe COVID-19 compared to no treatment.
- At least 8% of SARDs receiving oral outpatient treatment experienced COVID-19 rebound.

**How this study might affect research, practice, or policy?:**

- These results should encourage clinicians to prescribe and SARD patients to seek prompt outpatient COVID-19 treatment.
- This research provides an early estimate of the prevalence of COVID-19 rebound after oral outpatient treatment to quantify this risk to clinicians and SARD patients and encourage future research.

## INTRODUCTION

Outpatient SARS-CoV-2 treatment options include monoclonal antibodies, remdesivir, and oral medications such as nirmatrelvir/ritonavir and molnupiravir(1-4). For patients with systemic autoimmune rheumatic diseases (SARDs), effective COVID-19 treatments are important since altered immunity and immunosuppression may affect vaccine response(5, 6) and severity(7). COVID-19 rebound is a complication of oral outpatient antivirals, characterized by recurrence of symptoms and test positivity after regimen completion(8-12). However, there are limited data on outcomes with and without outpatient COVID-19 treatment among SARD patients, and the prevalence of rebound.

Therefore, we aimed to investigate outpatient SARS-CoV-2 treatment trends and outcomes, including rebound, in patients with SARDs. First, we examined temporal trends and the proportion of patients receiving outpatient treatment (monoclonal antibodies, oral medications, or outpatient remdesivir). Second, we compared severe COVID-19 outcomes for patients who did and did not receive outpatient treatment. Third, we described the prevalence of COVID-19 rebound among SARD patients who received oral outpatient treatment.

## METHODS

### Study population and design

We performed a retrospective cohort study investigating outpatient SARS-CoV-2 treatment and outcomes among SARD patients at Mass General Brigham (MGB). MGB is a multi-center healthcare system that includes a total of 14 hospitals as well as primary care and specialty outpatient centers in the greater Boston, Massachusetts area. We identified patients experiencing COVID-19 at MGB who were ≥18 years of age and had a SARD diagnosis. We studied SARD patients whose COVID-19 onset was between 23/Jan/2022 (when outpatient oral medications were first locally prescribed) to 30/May/2022. This study was approved by the MGB Institutional Review Board.

### Identification of COVID-19

We identified patients with COVID-19 using an electronic query of the MGB Research Patient Data Registry, which pulls data from the electronic health record (EHR). We identified COVID-19 as: 1) a positive SARS-CoV-2 polymerase chain reaction (PCR) or antigen test, and/or 2) a positive COVID-19 flag in the EHR. In MGB, a COVID-19 flag indicates a confirmed diagnosis of COVID-19 and captures patients with a positive test outside of our healthcare system. Patients are also flagged as having COVID-19 based on a positive home rapid antigen assay reported to providers or clinics and when ordering/administering outpatient treatments. In some cases, results from tests performed outside of MGB were pulled into the data warehouse. The index date was defined as the date of the first positive COVID-19 test/flag within the study dates.

### Identification of patients with SARDs

From this cohort of patients with COVID-19, we identified patients who had pre-existing SARDs at the onset of COVID-19. We previously described identification of SARDs at MGB using administrative data for COVID-19 studies in detail, validated with 90% positive predictive value(13). Briefly, SARDs fulfill these two criteria: 1) ≥2 International Classification of Diseases (ICD)-10 codes (see **Supplementary Table 1**) for a SARD within two years prior to the index date and separated by ≥30 days, and 2) prescription of a disease-modifying antirheumatic drug [DMARD] within 12 months prior to the index date and/or a prescription for systemic glucocorticoid within 6 months of the index date (see **Supplementary Table 2**). We required all patients to have received immunomodulatory medications to enhance specificity. Patients with osteoarthritis, fibromyalgia, or crystalline arthritis without another concomitant SARD diagnosis were not included since these conditions are generally not treated with long-term immunomodulators and are often managed by non-rheumatologists.

### Exposure variable: Any vs. no outpatient SARS-CoV-2 treatment

The primary exposure of the study was any vs. no outpatient treatment for SARS-CoV-2. Secondary exposures were specific treatments among those with adequate sample size. Since a pre-specified aim of our study was to investigate oral outpatient treatment options, we only analyzed a time period when these were available locally. Nirmatrelvir/ritonavir received emergency authorization to high-risk individuals from the US Food and Drug Administration on 22/Dec/2021(14); molnupiravir received authorization on 23/Dec/2021(15). The start of this study was 23/Jan/2022, when these medications were first prescribed locally.

We performed manual medical record review of all identified SARD patients with COVID-19 during the study period to accurately classify outpatient treatments or verify lack of outpatient treatment. If a patient received more than one therapy in the outpatient setting, they were classified as combination treatment.

### Outcome: Severe COVID-19

The primary outcome was severe COVID-19, defined as hospitalization and/or death within 30 days after index date. This was identified using electronic query, as in our previous studies(16-20). In a sensitivity analysis, we required that the outcome occur at least 1 day after the index date, since some may have been unable to receive outpatient treatment in time to prevent hospitalization(21). Also, some may have been incidentally found to have COVID-19 while hospitalized for other reasons and therefore ineligible for outpatient therapy.

### COVID-19 rebound after oral outpatient SARS-CoV-2 treatment

Among patients who received oral outpatient SARS-CoV-2 treatment, we performed medical record review to identify those who experienced COVID-19 rebound documented in the EHR. As in a previous study(11), COVID-19 rebound was defined as: 1) completion of oral SARS-CoV-2 oral outpatient regimen, 2) documentation of negative then newly-positive SARS-CoV-2 tests within 7 days of completion, and 3) recurrence in COVID-19 symptoms after improvement in most or all symptoms within 7 days of completion. Patients who had little or no improvement in symptoms throughout follow-up were not considered as rebound cases. Similarly, patients with prolonged viral shedding(22) without a negative test in the interim were not considered as rebound cases.

### SARD characteristics

We classified each patient’s primary SARD diagnosis using ICD-10 codes, as described previously(13). Immunomodulatory medications were identified using prescription data preceding the index date. For conventional and biologic synthetic DMARDs, we separately considered the most recent prescription(s). For CD20 inhibitors, we classified exposure if last received within one year before the index date due to lengthy effects that can impact COVID-19 vaccine response and COVID-19 severity(19, 23, 24).

### Other covariates

We used electronic query to identify most covariates. Demographic factors were: age, sex, race (White, Black, Asian, other, unknown), and Hispanic ethnicity. Lifestyle factors were: body mass index (continuous) and smoking status (never, past, current, missing). We calculated the Charlson Comorbidity Index (CCI)(25) from ICD-10 codes in the one year prior to index date. We also identified individual components of the CCI as well as interstitial lung disease, which has been previously associated with worse COVID-19 outcomes(26). The estimated glomerular filtration rate (eGFR) was calculated using the CKD-EPI equation without the race multiplier(27). We categorized eGFR as <30 (severe), 30 to <60 (moderate), or ≥60 (normal) mL/min/1.73 m^2^ since this impacts nirmatrelvir/ritonavir dosing and eligibility(28).

COVID-19 vaccine types and dates were extracted from the EHR. As in our prior study(18), vaccination status was classified as unvaccinated, partially vaccinated, 2 doses of mRNA or 1 dose of adenovirus vaccines, or additional vaccine doses. For patients initially classified as unvaccinated or partially vaccinated, we performed medical record review to accurately classify patients who may have received vaccines outside of Massachusetts. For patients who had received vaccines, we also classified whether their most recent vaccine dose before index date was < or ≥6 months, since humoral immunity wanes(29, 30). We used electronic query to identify previous COVID-19 prior to the current episode. We performed manual medical record review to determine tixagevimab/cilgavimab use(31).

### Statistical analysis

We plotted the total number of COVID-19 cases per calendar week in the study period and subdivided this by outpatient treatment status (nirmatrelvir/ritonavir, molnupiravir, monoclonal antibodies, remdesivir, combination, or untreated as outpatient). We calculated the p for trend of the proportion treated across the ordinal variable of calendar week. We reported baseline characteristics of the entire study sample using descriptive statistics according to outpatient treatment exposure status.

The primary analysis compared any vs. no outpatient SARS-CoV-2 treatment for the outcome of severe COVID-19. We first performed an unadjusted logistic regression model to calculate the odds ratios (ORs) and 95% confidence intervals (CIs) for severe COVID-19. In the multivariable model, we included age, CCI, eGFR, and race (White vs. non-White) as possible confounders associated with outpatient treatment and severe COVID-19. We did not include vaccination status since there were few unvaccinated patients.

We performed similar analyses for other comparisons of outpatient treatments for risk of severe COVID-19. These included nirmatrelvir/ritonavir vs. no outpatient treatment, monoclonal antibodies vs. no outpatient treatment, nirmatrelvir/ritonavir vs. all others, monoclonal antibodies vs. all others, nirmatrelvir/ritonavir vs. monoclonal antibodies. We were unable to investigate molnupiravir, outpatient remdesivir, or combination use since there were few patients that received these therapies. In a sensitivity analysis, we only considered severe COVID-19 outcomes that occurred at least one day after the index date.

We also performed subgroup analyses for COVID-19 severity for the following comparisons that each had adequate sample size: any vs. no outpatient treatment, nirmatrelvir/ritonavir vs. no outpatient treatment, and monoclonal antibodies vs. no outpatient treatment. We investigated the following subgroups: age (<65, ≥65 years), sex (male, female), dichotomized CCI (0 to 1, ≥2), eGFR (<30, ≥30 mL/min/1.73 m^2^), vaccination status (unvaccinated, 2 mRNA or 1 adenovirus, or additional doses [none were partially vaccinated]), and duration since last vaccine dose (_≤_6 months, >6 months). We reported the numbers of outcomes and total n in each subgroup and multivariable ORs and 95%CIs in forest plots.

For COVID-19 rebound, we reported the number of confirmed cases over the denominator of patients who received either nirmatrelvir/ritonavir or molnupiravir (either as monotherapy or in combination with other medications such as monoclonal antibodies). We reported descriptive statistics of baseline characteristics of these patients. We did not perform association analyses for COVID-19 rebound since some patients may have experienced COVID-19 rebound but may not have been documented in the EHR.

We considered a two-sided p value of <0.05 as statistically significant in all analyses. All analyses were performed using SAS v.9.4 (Cary, NC).

## RESULTS

### Study sample and temporal trends of outpatient treatment

We identified 704 SARD patients with COVID-19 between 23/Jan/2022 and 30/May/2022. **Figure 1** shows the number of COVID-19 cases and their outpatient treatments among SARDs over the study period. The proportion treated as outpatient increased over calendar time; 35% were treated at the start of the study compared to 65% at the study end; p for trend <0.001).

**Figure 1.**
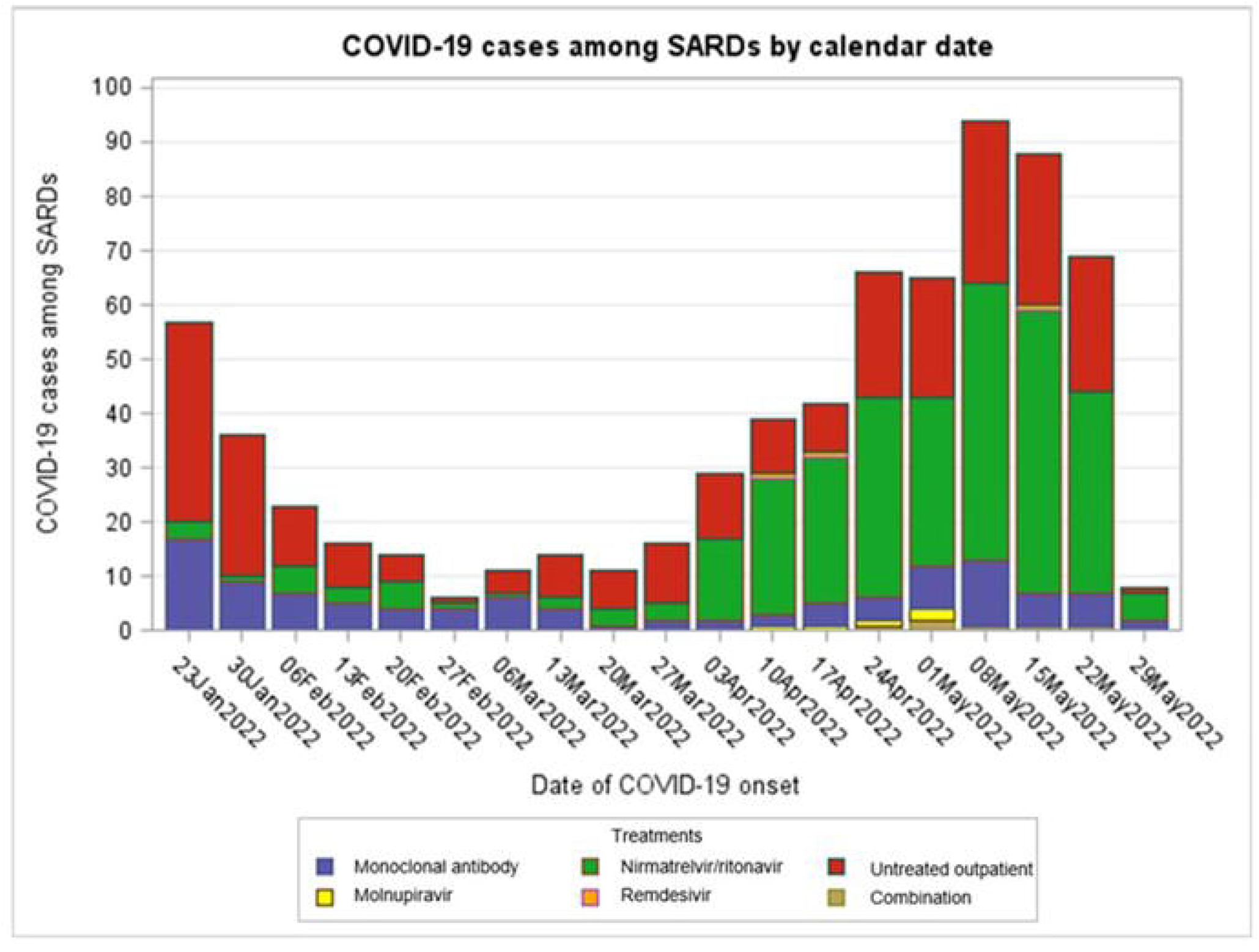
COVID-19 cases over calendar time among patients with systemic autoimmune rheumatic diseases by outpatient SARS-CoV-2 treatments (n=704). Note that the week of 29/May/2022 only includes two days. COVID-19, coronavirus disease 2019; SARD, systemic autoimmune rheumatic disease; SARS-CoV-2, severe acute respiratory syndrome coronavirus 2.

### Outpatient SARS-CoV-2 treatments and baseline characteristics

A total of 426 (61%) patients received any outpatient SARS-CoV-2 treatment: 307 (44%) with nirmatrelvir/ritonavir, 105 (15%) with monoclonal antibodies, 5 (0.7%) with molnupiravir, 3 (0.4%) with outpatient remdesivir, and 6 (0.9%) with combinations (n=4 nirmatrelvir/ritonavir and monoclonal antibodies; n=2 molnupiravir and monoclonal antibodies). A total of 278 (39%) received no outpatient SARS-CoV-2 treatment (**Table 1**).

**Table 1.** Demographics, lifestyle, comorbidities, and previous COVID-19 characteristics of SARD patients at COVID-19 onset by outpatient SARS-CoV-2 treatment (n=704).

| Characteristic | All SARD patients with COVID-19 (n=704) | Outpatient treatment |  |  | No treatment (n=278) |
| --- | --- | --- | --- | --- | --- |
|  |  | Any treatment (n=426)* | Nirmatrelvir/ritonavir (n=307) | Monoclonal antibody (n=105) |  |
| Demographics |  |  |  |  |  |
| Mean age (SD), years | 58.4 (15.9) | 58.3 (15.6) | 57.1 (14.9) | 61.2 (17.5) | 58.7 (16.4) |
| Female | 536 (76.4%) | 331 (77.7%) | 235 (76.6%) | 83 (80.0%) | 205 (73.7%) |
| Race |  |  |  |  |  |
| Asian | 20 (2.8%) | 11 (2.6%) | 9 (2.9%) | 1 (1.0%) | 9 (3.2%) |
| Black or African American | 39 (5.5%) | 19 (4.5%) | 15 (4.9%) | 4 (3.8%) | 20 (7.2%) |
| Other | 36 (5.1%) | 20 (4.7%) | 16 (5.2%) | 4 (3.8%) | 16 (5.8%) |
| White | 590 (83.8%) | 367 (86.2%) | 259 (84.4%) | 95 (90.5%) | 223 (80.2%) |
| Unknown | 19 (2.7%) | 9 (2.1%) | 8 (2.6%) | 1 (1.0%) | 10 (3.6%) |
| Hispanic or Latinx ethnicity | 7 (1.0%) | 5 (1.2%) | 4 (1.3%) | 1 (1.0%) | 2 (1.0%) |
| Lifestyle |  |  |  |  |  |
| Mean body mass index (SD), kg/m<sup>2</sup> | 27.7 (7.8) | 28.1 (7.4) | 27.7 (7.3) | 28.9 (7.4) | 27.0 (8.3) |
| Smoking status |  |  |  |  |  |
| Never | 412 (58.5%) | 260 (61.0%) | 202 (65.8%) | 50 (47.6%) | 152 (54.7%) |
| Past | 244 (34.7%) | 146 (34.7%) | 92 (30.0%) | 48 (45.7%) | 98 (35.4%) |
| Current | 31 (4.4%) | 15 (3.5%) | 9 (2.9%) | 6 (5.7%) | 16 (5.8%) |
| Unknown | 17 (2.4%) | 5 (1.2%) | 4 (1.3%) | 1 (1.0%) | 12 (4.3%) |
| Comorbidities |  |  |  |  |  |
| Median Charlson Comorbidity Index (IQR) | 1 (1, 3) | 1 (1, 3) | 1 (1, 2) | 2 (1, 5) | 2 (1, 4) |
| Charlson Comorbidity Index categories |  |  |  |  |  |
| 0 | 100 (14.2%) | 56 (13.2%) | 47 (15.3%) | 8 (7.6%) | 44 (15.8%) |
| 1 | 272 (38.6%) | 180 (42.3%) | 151 (49.2%) | 25 (23.8%) | 92 (33.1%) |
| 2 | 116 (16.5%) | 75 (17.6%) | 51 (16.6%) | 22 (21.0%) | 41 (14.8%) |
| ≥3 | 216 (30.7%) | 115 (27.0%) | 58 (18.9%) | 50 (47.6%) | 101 (36.3%) |
| Individual comorbidities |  |  |  |  |  |
| Hypertension | 301 (42.8%) | 167 (39.2%) | 103 (33.6%) | 57 (54.3%) | 134 (48.2%) |
| Asthma | 112 (15.9%) | 68 (16.0%) | 43 (14.0%) | 22 (21.0%) | 44 (15.8%) |
| Cancer excluding non-melanoma skin cancer | 106 (15.1%) | 57 (13.4%) | 34 (11.1%) | 21 (20.0%) | 49 (17.6%) |
| Coronary artery disease | 98 (13.9%) | 53 (12.4%) | 23 (7.5%) | 26 (24.8%) | 45 (16.2%) |
| Chronic kidney disease | 95 (13.5%) | 51 (12.0%) | 28 (9.0%) | 20 (19.1%) | 44 (15.8%) |
| Diabetes | 90 (12.8%) | 43 (10.1%) | 19 (6.2%) | 19 (18.1%) | 47 (16.9%) |
| Heart failure | 64 (9.1%) | 26 (6.1%) | 4 (1.3%) | 19 (18.1%) | 38 (13.7%) |
| Chronic obstructive pulmonary disease | 44 (6.3%) | 20 (4.7%) | 4 (1.3%) | 15 (14.3%) | 24 (8.6%) |
| Interstitial lung disease | 42 (6.0%) | 20 (4.7%) | 9 (2.9%) | 11 (10.5%) | 22 (7.9%) |
| Median eGFR (IQR), mL/min/1.73 m<sup>2</sup> | 87 (71, 100.5) | 86 (71, 101) | 88 (77, 101) | 80 (64, 97) | 87.5 (70, 100) |
| Categorical eGFR, mL/min/1.73 m<sup>2</sup> |  |  |  |  |  |
| ≥60 | 614 (87.2%) | 378 (88.7%) | 284 (92.5%) | 86 (81.9%) | 236 (84.9%) |
| ≥30 to <60 | 75 (10.6%) | 42 (9.9%) | 23 (7.5%) | 14 (13.3%) | 33 (11.9%) |
| <30 | 15 (2.1%) | 6 (1.4%) | 0 (0.00%) | 5 (4.8%) | 9 (3.2%) |
| Previous COVID-19 immunity |  |  |  |  |  |
| Vaccination status |  |  |  |  |  |
| Unvaccinated | 27 (3.8%) | 9 (2.1%) | 8 (2.6%) | 1 (1.0%) | 18 (6.5%) |
| Partially vaccinated | 0 (0.0%) | 0 (0.0%) | 0 (0.0%) | 0 (0.0%) | 0 (0.0%) |
| 2 doses mRNA or 1 dose adenovirus | 112 (15.9%) | 56 (13.2%) | 41 (13.4%) | 13 (12.4%) | 56 (20.1%) |
| Additional doses | 565 (80.3%) | 361 (84.7%) | 258 (84.0%) | 91 (86.7%) | 204 (73.4%) |
| Tixagevimab/cilgavimab use | 12 (1.7%) | 8 (1.9%) | 5 (1.6%) | 3 (1.9%) | 4 (1.4%) |
| Previous COVID-19 infection | 10 (1.4%) | 8 (1.8%) | 8 (2.6%) | 0 (0.0%) | 2 (0.7%) |

\*Characteristics of other outpatient treatments: molnupiravir use (n=5), remdesivir use (n=3), and combination use (n=6; 4 received nirmatrelvir/ritonavir and monoclonal antibodies and 2 received molnupiravir and monoclonal antibodies) are shown in **Supplemental Table 3**.
COVID-19, coronavirus disease 2019; eGFR, estimated glomerular filtration rate; IQR, interquartile range; SARD, systemic autoimmune rheumatic disease; SARS-CoV-2, severe acute respiratory syndrome coronavirus 2; SD, standard deviation.

The mean age was 58.4 years, 76% were female, 84% were White, and 96% were vaccinated. Those who received outpatient treatment vs. none were more often female (77.7% vs. 73.7%) and White (86.5% vs. 80.2%), less likely to have severe kidney impairment (1.4% vs. 3.2%), and less likely to be unvaccinated (2.1% vs. 6.5%).

SARD characteristics are shown in **Table 2**. 49.2% had rheumatoid arthritis, 16.1% had psoriatic arthritis, and 12.4% had systemic lupus. Conventional synthetic DMARDs were used in 68.8%, most frequently methotrexate (38.5%) and hydroxychloroquine (30.4%). Biologic DMARDs were used in 36.7%, most frequently tumor necrosis factor inhibitors (20.5%). Characteristics of patients that used other outpatient treatments are in **Supplemental Table 3**.

**Table 2.** Rheumatic disease characteristics of SARD patients at COVID-19 onset by outpatient SARS-CoV-2 treatment (n=704).

| Characteristic | All SARD patients with COVID-19 (n=704) | Outpatient treatment |  |  | No outpatient treatment (n=278) |
| --- | --- | --- | --- | --- | --- |
|  |  | Any treatment (n=426)* | Nirmatrelvir/ritonavir (n=307) | Monoclonal antibodies (n=105) |  |
| **Rheumatic disease diagnosis** |  |  |  |  |  |
| Rheumatoid arthritis | 347 (49.3%) | 212 (50.8%) | 156 (50.8%) | 52 (49.5%) | 135 (48.6%) |
| Psoriatic arthritis | 113 (16.1%) | 72 (16.9%) | 62 (20.2%) | 9 (8.6%) | 41 (14.8%) |
| Systemic lupus erythematosus | 87 (12.4%) | 54 (12.7%) | 37 (12.1%) | 13 (12.4%) | 33 (11.9%) |
| Giant cell arteritis and/or polymyalgia rheumatica | 45 (6.4%) | 28 (6.6%) | 13 (4.2%) | 14 (13.3%) | 17 (6.1%) |
| Sjogren's syndrome | 24 (3.4%) | 12 (2.8%) | 7 (2.3%) | 4 (3.8%) | 12 (4.3%) |
| ANCA-associated vasculitis and other miscellaneous vasculitis | 20 (2.8%) | 15 (3.5%) | 9 (2.9%) | 5 (4.8%) | 5 (1.8%) |
| Systemic sclerosis | 17 (2.4%) | 6 (1.4%) | 4 (1.3%) | 2 (1.9%) | 11 (3.9%) |
| Axial spondyloarthritis | 11 (1.6%) | 8 (1.9%) | 5 (1.6%) | 2 (1.9%) | 3 (1.1%) |
| Mixed connective tissue disease | 11 (1.6%) | 5 (1.2%) | 2 (0.7%) | 2 (1.9%) | 6 (2.2%) |
| Antiphospholipid antibody syndrome | 6 (0.9%) | 3 (0.7%) | 3 (1.0%) | 0 (0.0%) | 3 (1.1%) |
| Behcet disease | 6 (0.9%) | 3 (0.7%) | 3 (1.0%) | 0 (0.0%) | 3 (1.9%) |
| Takayasu arteritis | 3 (0.4%) | 2 (0.5%) | 1 (0.3%) | 1 (1.0%) | 1 (0.4%) |
| Idiopathic inflammatory myositis | 1 (0.1%) | 0 (0.0%) | 0 (0.0%) | 0 (0.0%) | 1 (0.4%) |
| Juvenile idiopathic arthritis | 1 (0.1%) | 1 (0.2%) | 1 (0.3%) | 0 (0.0%) | 0 (0.0%) |
| Multiple primary rheumatic diseases | 12 (1.7%) | 5 (1.2%) | 4 (1.3%) | 1 (1.0%) | 7 (2.5%) |
| **Immunomodulatory medications** |  |  |  |  |  |
| Oral glucocorticoid | 51 (7.2%) | 18 (4.2%) | 6 (2.0%) | 12 (11.4%) | 33 (11.9%) |
| Conventional synthetic DMARDs | 484 (68.8%) | 284 (66.7%) | 205 (66.8%) | 71 (67.6%) | 200 (72%) |
| Methotrexate | 232 (33.0%) | 138 (32.4%) | 106 (34.5%) | 32 (30.5%) | 94 (33.8%) |
| Hydroxychloroquine | 214 (30.4%) | 117 (27.5%) | 84 (27.4%) | 26 (24.8%) | 97 (34.9%) |
| Mycophenolate mofetil/mycophenolic acid | 47 (6.7%) | 27 (6.3%) | 14 (4.6%) | 12 (11.4%) | 20 (7.2%) |
| Leflunomide | 42 (6.0%) | 31 (7.3%) | 23 (7.5%) | 7 (6.7%) | 11 (4.0%) |
| Sulfasalazine | 35 (5.0%) | 18 (4.2%) | 15 (4.9%) | 3 (2.9%) | 17 (6.1%) |
| Tacrolimus | 34 (4.8%) | 22 (5.2%) | 9 (2.9%) | 12 (11.4%) | 12 (4.3%) |
| Azathioprine | 23 (3.3%) | 14 (3.3%) | 8 (2.6%) | 5 (4.8%) | 9 (3.2%) |
| Cyclosporine | 17 (2.4%) | 9 (2.1%) | 6 (2.0%) | 2 (2.0%) | 8 (2.9%) |
| Cyclophosphamide | 5 (0.7%) | 4 (0.9%) | 4 (1.3%) | 0 (0.0%) | 1 (0.4%) |
| Apremilast | 3 (0.4%) | 3 (0.7%) | 2 (0.7%) | 1 (1.0%) | 0 (0.0%) |
| Biologic DMARDs | 258 (36.7%) | 180 (42.3%) | 133 (43.3%) | 39 (37.1%) | 78 (28.1%) |
| TNF inhibitor | 144 (20.5%) | 104 (24.4%) | 86 (28.0%) | 15 (14.3%) | 40 (14.4%) |
| CD20 inhibitor | 38 (5.4%) | 26 (6.1%) | 13 (4.2%) | 10 (9.5%) | 12 (4.3%) |
| IL-6 receptor inhibitor | 22 (3.1%) | 17 (4.0%) | 10 (3.3%) | 5 (4.8%) | 5 (1.8%) |
| IL-17 inhibitor | 20 (2.8%) | 14 (3.3%) | 13 (4.2%) | 1 (1.0%) | 6 (2.2%) |
| CTLA-4 immunoglobulin | 20 (2.8%) | 11 (2.6%) | 8 (2.6%) | 3 (2.9%) | 9 (3.2%) |
| IL-23 inhibitor | 5 (0.7%) | 2 (0.5%) | 1 (0.3%) | 1 (1.0%) | 3 (1.1%) |
| B-cell activating factor inhibitor | 4 (0.6%) | 3 (0.7%) | 1 (0.3%) | 2 (1.9%) | 1 (0.4%) |
| IL-12/IL-23 inhibitor | 3 (0.4%) | 2 (0.5%) | 1 (0.3%) | 1 (1.0%) | 1 (0.4%) |
| IL-5 inhibitor | 3 (0.4%) | 1 (0.2%) | 0 (0.0%) | 1 (1.0%) | 2 (0.7%) |
| IL-1 inhibitor | 2 (0.3%) | 2 (0.5%) | 1 (0.3%) | 1 (1.0%) | 0 (0.0%) |
| Targeted synthetic DMARD |  |  |  |  |  |
| JAK inhibitor | 16 (2.3%) | 11 (2.6%) | 8 (2.6%) | 3 (2.9%) | 5 (1.8%) |

\*Characteristics of other outpatient treatments: molnupiravir use (n=5), remdesivir use (n=3), and combination use (n=6; 4 received nirmatrelvir/ritonavir and monoclonal antibodies and 2 received molnupiravir and monoclonal antibodies) are shown in **Supplemental Table 3**.
ANCA, antineutrophil cytoplasmic antibodies; CTLA-4, cytotoxic T-lymphocyte-associated protein 4; DMARDs, disease-modifying antirheumatic drugs; IL, interleukin; JAK, Janus kinase; SARD, systemic autoimmune rheumatic disease; SARS-CoV-2, severe acute respiratory syndrome coronavirus 2; TNF, tumor necrosis factor.

### Severe COVID-19 outcomes

A total of 58 (8.2%) hospitalizations and 3 (0.4%) deaths occurred within 30 days of COVID-19 onset (**Table 3**). The composite primary outcome of hospitalization/death occurred in 58 (8.2%). Of the 426 patients treated as an outpatient, nine (2.1%, 1 death) had severe COVID-19 compared to 49 (17.6%, 2 deaths) of 278 untreated patients. Of the 307 patients treated with nirmatrelvir/ritonavir, four (1.3%, 1 death) had severe COVID-19 outcomes. Of the 105 patients treated with monoclonal antibodies, five (4.8%) had hospitalizations. No severe COVID-19 outcomes occurred among the 5 patients who received molnupiravir, outpatient remdesivir, or combinations.

**Table 3.** COVID-19 outcomes by outpatient SARS-CoV-2 treatment among SARD patients (n=704).

| **Outcome** | **All SARD patients with COVID-19 (n=704)** | **Any outpatient treatment (n=426)\*** | **Nirmatrelvir/ritonavir use (n=307)** | **Monoclonal antibody use (n=105)** | **No outpatient treatment (n=278)** |
| --- | --- | --- | --- | --- | --- |
| Hospitalization | 58 (8.2%) | 9 (2.1%) | 4 (1.3%) | 5 (4.8%) | 49 (17.6%) |
| Death | 3 (0.4%) | 1 (0.2%) | 1 (0.3%) | 0 (0.0%) | 2 (0.7%) |
| Severe COVID-19 (hospitalization or death) | 58 (8.2%) | 9 (2.1%) | 4 (1.3%) | 5 (4.8%) | 49 (17.6%) |
| Rebound | N/A | N/A | 25/311** (8.0%) | N/A | N/A |

\*There were no severe COVID-19 outcomes among molnupiravir (n=5), remdesivir (n=3), or combination (n=6; 4 received nirmatrelvir/ritonavir and monoclonal antibodies and 2 received molnupiravir and monoclonal antibodies) users.
\*\*The denominator for COVID-19 rebound also includes 4 patients who used nirmatrelvir/ritonavir as a combination with monoclonal antibodies. There was also 1 COVID-19 rebound case among 7 molnupiravir users (14.3%).
COVID-19, coronavirus disease 2019; N/A, not applicable; SARD, systemic autoimmune rheumatic disease; SARS-CoV-2, severe acute respiratory syndrome coronavirus 2.

The majority of patients who received or did not receive treatment were previously vaccined (97.9% vs. 93.5%, respectively). Among the 27 unvaccinated patients, there were 2 severe COVID-19 outcomes; neither of these received outpatient treatment.

### Severe COVID-19 risk by outpatient treatment

Results comparing outpatient treatments for risk of severe COVID-19 are shown in **Table 4**. After adjustment for age, CCI, eGFR, and race, any outpatient treatment had an adjusted OR (aOR) for severe COVID-19 of 0.12 (95%CI 0.05 to 0.25) compared to no outpatient treatment.

**Table 4.** Odds ratios for severe COVID-19 (hospitalization or death) by outpatient SARS-CoV-2 treatment status.

| **Comparisons (reference=second group listed)** | **Unadjusted OR for severe COVID-19 (95%CI)** | **Multivariable\* OR for severe COVID-19 (95%CI)** |
| --- | --- | --- |
| **Primary analysis** |  |  |
| Any outpatient treatment vs. no outpatient treatment | 0.10 (0.05, 0.21) | 0.12 (0.05, 0.25) |
| **Secondary analyses** |  |  |
| Nirmatrelvir/ritonavir vs. no outpatient treatment | 0.06 (0.02, 0.17) | 0.08 (0.03, 0.24) |
| Monoclonal antibodies vs. no outpatient treatment | 0.23 (0.09, 0.60) | 0.20 (0.07, 0.54) |
| Nirmatrelvir/ritonavir vs. all others | 0.08 (0.03, 0.23) | 0.12 (0.04, 0.34) |
| Monoclonal antibodies vs. all others | 0.52 (0.20, 1.32) | 0.35 (0.13, 0.97) |
| Nirmatrelvir/ritonavir vs. monoclonal antibodies | 0.26 (0.07, 1.00) | 0.46 (0.11, 1.97) |

\*Adjusted for continuous age, continuous Charlson Comorbidity Index, continuous estimated glomerular filtration rate, and race.
CI, confidence interval; COVID-19, coronavirus disease 2019; OR, odds ratio; SARS-CoV-2, severe acute respiratory syndrome coronavirus 2.

In the secondary analyses, nirmatrelvir/ritonavir (aOR 0.08, 95%CI 0.03 to 0.24) and monoclonal antibodies (aOR 0.20, 95%CI 0.07 to 0.54) were each associated with lower odds of severe COVID-19 compared to no treatment. Comparing COVID-19 outcomes among nirmatrelvir/ritonavir vs. monoclonal antibodies, there was no statistical difference (aOR 0.46, 95%CI 0.11 to 1.97).

### Severe COVID-19 risk among subgroups

**Figure 2** shows forest plots investigating odds of severe COVID-19 with and without outpatient treatment among these subgroups: age (<65, ≥65 years), sex (male, female), CCI (0 to 1, ≥2), eGFR (≥30, <30 mL/min/1.73 m^2^), vaccination status (unvaccinated, 2 mRNA or 1 adenovirus, additional doses), and duration since last vaccine dose (_≤_6 months, >6 months). **Figure 2A** shows any vs. no outpatient treatment, **Figure 2B** shows nirmatrelvir/ritonavir vs. no outpatient treatment, and **Figure 2C** shows monoclonal antibodies vs. no outpatient treatment. Findings from our primary analysis remained robust across all subgroups.

**Figure 2.**
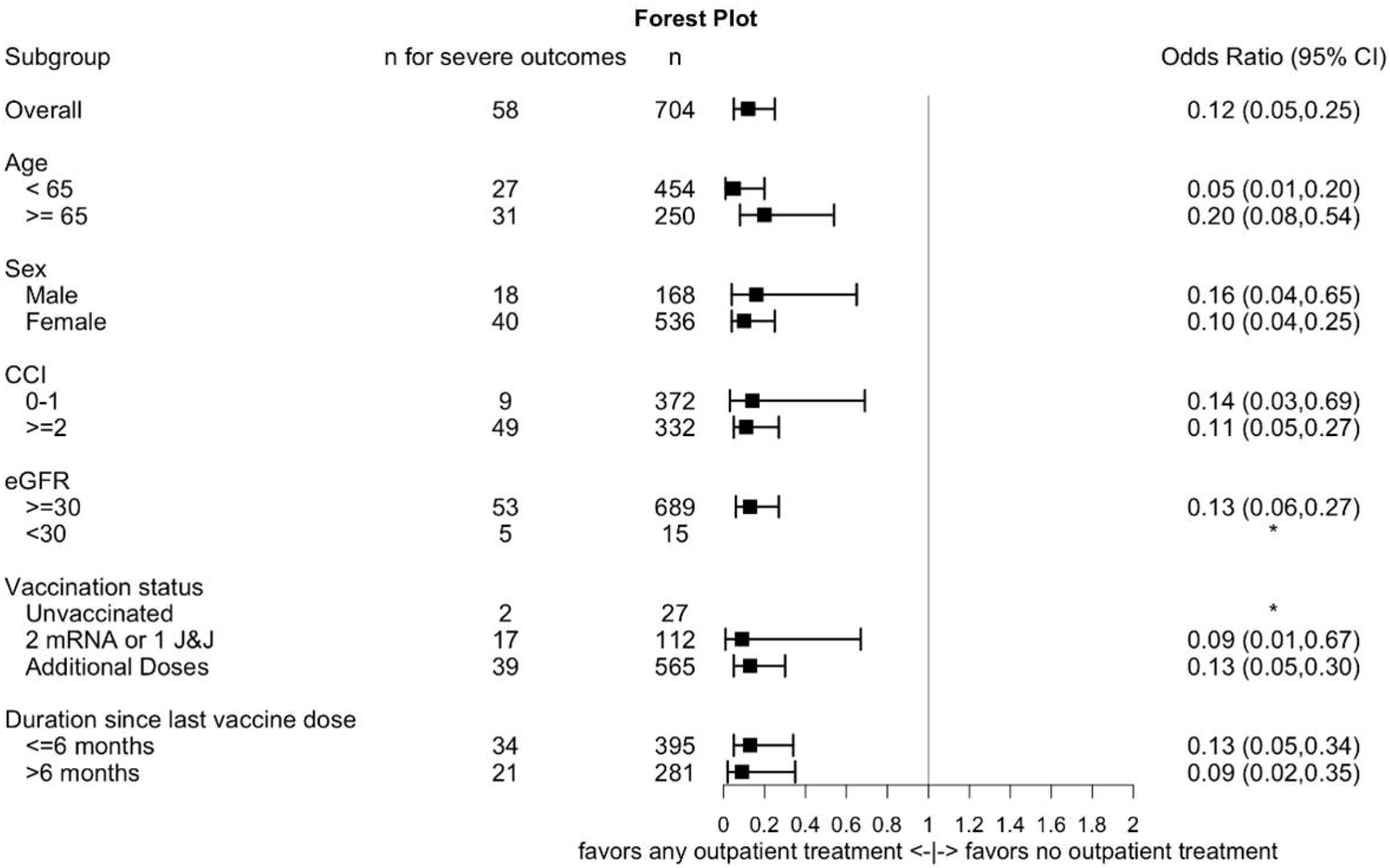
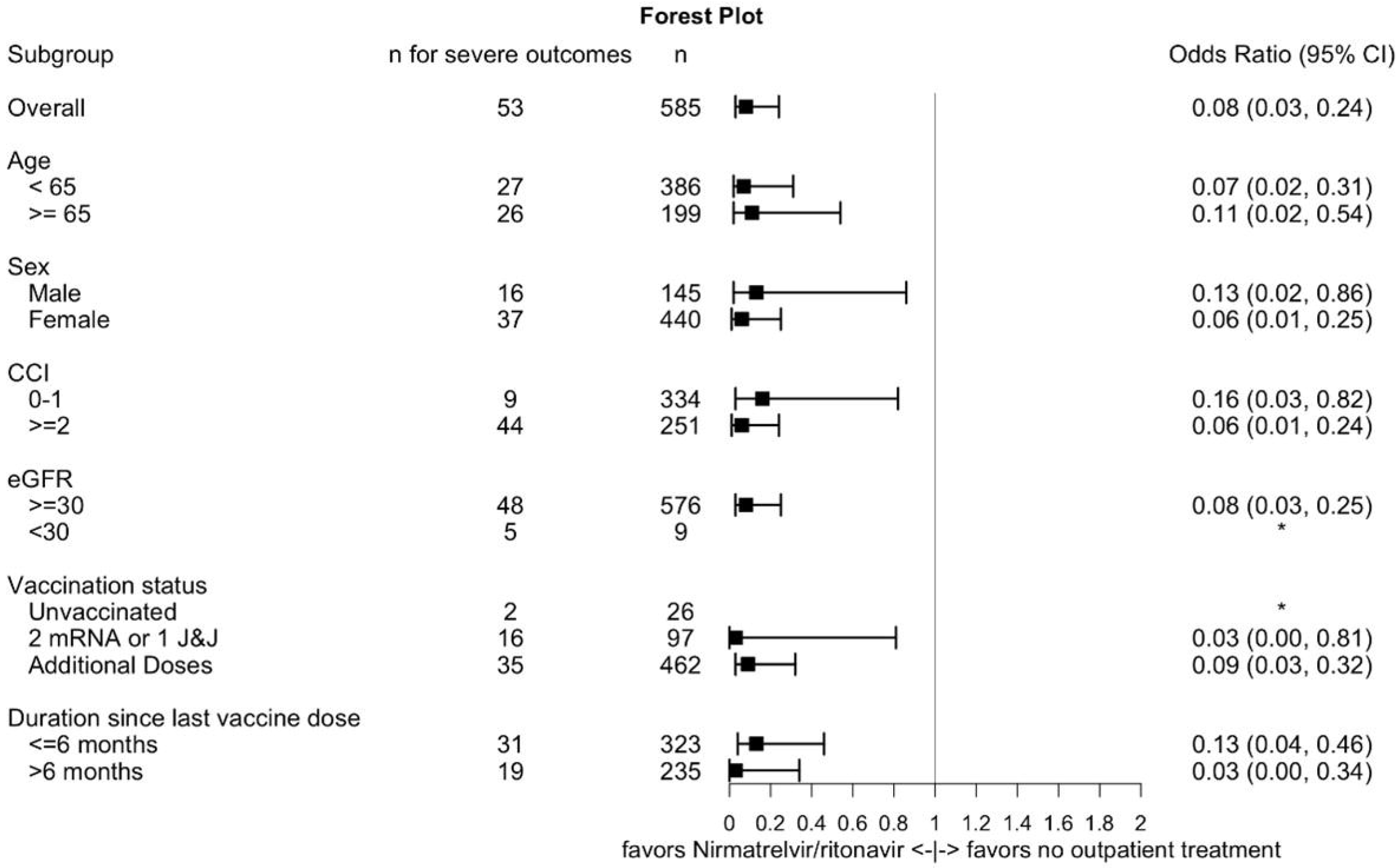
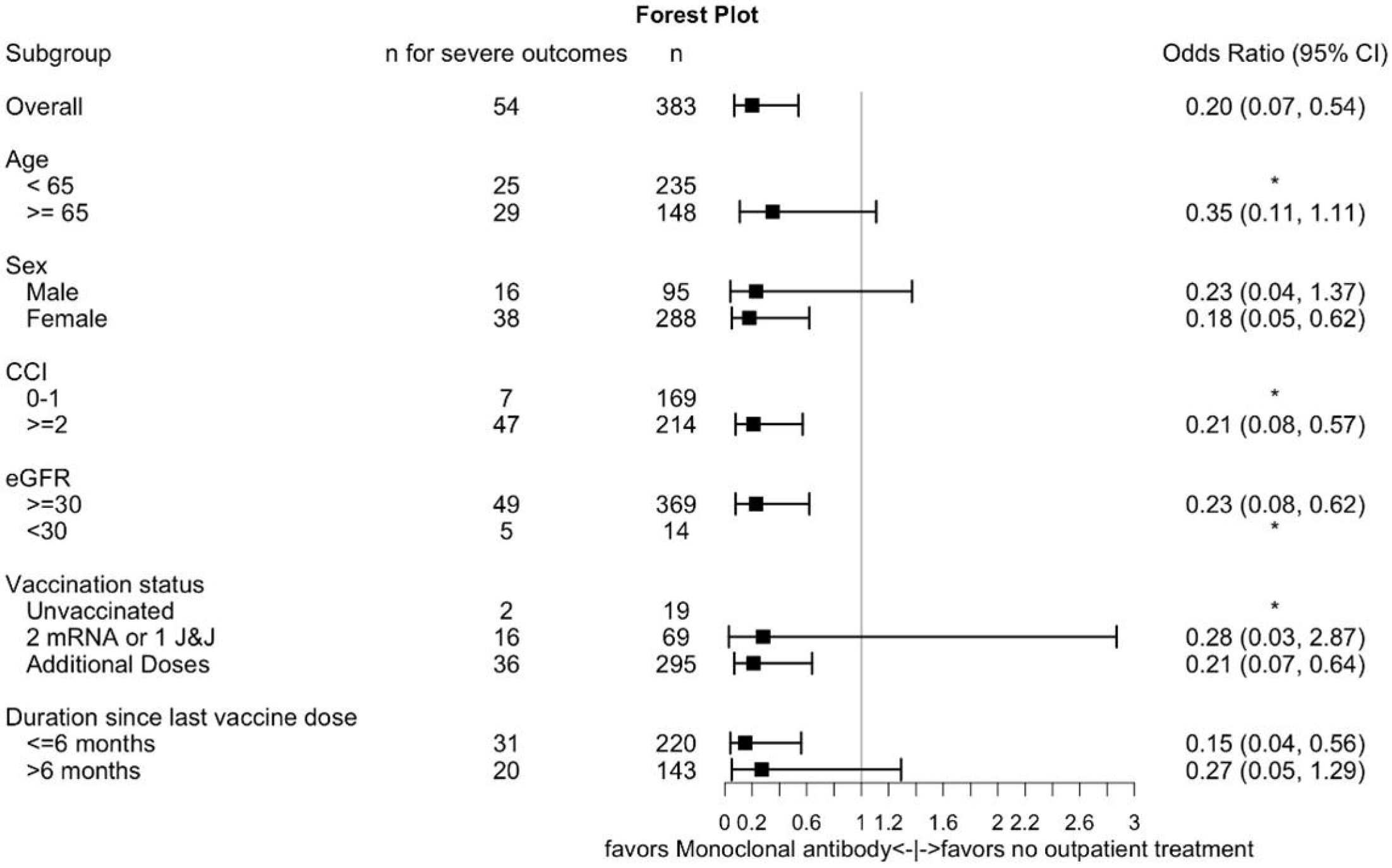
Forest plots of subgroups for odds of severe COVID-19 (hospitalization or death) comparing A) any vs. no outpatient treatment, B) nirmatrelvir/ritonavir vs. no outpatient treatment, and C) monoclonal antibodies vs. no outpatient treatment. Adjusted for continuous age, continuous Charlson Comorbidity Index, continuous estimated glomerular filtration rate, and race. *Model did not converge due to few outcomes.

### Sensitivity analysis for severe COVID-19

In the sensitivity analysis that considered only severe COVID-19 occurring at least one day after the index date, there were a total of 38 (5.4%) outcomes (2.1% among any outpatient treatment vs. 10.4% without outpatient treatment, **Supplemental Table 4**). Any outpatient treatment had an aOR for severe COVID-19 of 0.22 (95%CI 0.10 to 0.48) compared to no outpatient treatment (**Supplemental Table 5**).

### COVID-19 rebound

We identified 25 (8%) of 318 who received oral outpatient SARS-CoV-2 treatments and had documented COVID-19 rebound. Among nirmatrelvir/ritonavir users, 24/311(8%) had COVID-19 rebound. Among molnupiravir users, 1/7(14%) had COVID-19 rebound. Characteristics of those that experienced COVID-19 rebound are in **Supplemental Table 6**.

## DISCUSSION

In this contemporary cohort of SARD patients with COVID-19, outpatient treatment with antivirals or monoclonal antibodies was associated with 88% lower odds of severe COVID-19 compared to no outpatient treatment. Outpatient COVID-19 treatment increased over the study period; the most common outpatient treatments were nirmatrelvir/ritonavir and monoclonal antibodies. Among those who received oral outpatient treatment, the prevalence of COVID-19 rebound was 8%, a likely conservative estimate due to the requirement of EHR documentation. These findings highlight the importance of early outpatient treatment in this vulnerable population, even among those vaccinated, and emphasize the need to further investigate COVID-19 rebound in SARDs.

Despite advances in prevention and associated improvements in outcomes, patients with SARDs remain at elevated risk for SARS-CoV-2 infection, severe outcomes, and prolonged symptom duration, especially those who are on B cell depleting therapy or have comorbid conditions like interstitial lung disease(19, 24, 26). Although vaccination reduces risk for severe outcomes, RA patients have elevated risks for SARS-CoV-2 infection and severe outcomes compared to the general population(32). Thus, even with improving COVID-19 outcomes, some SARD patients remain vulnerable to poor COVID-19 outcomes(33, 34). In this context, our novel findings regarding the 80% lower odds of severe COVID-19 associated with outpatient treatment are an important reminder to clinicians to consider early outpatient treatment for SARD patients with COVID-19. Importantly, our findings persisted across all subgroups examined, including younger patients and those who remained unvaccinated during a study period characterized by predominance of the highly contagious Omicron variants. The majority of treated patients in our study received nirmatrelvir/ritonavir and monoclonal antibodies; few received molnupiravir (1.2%), outpatient remdesivir (0.7%), or combinations (1.4%). Whether similar patterns of use and benefit will be observed in other centers or with use of these less frequently used treatments requires further investigation.

COVID-19 rebound is characterized by re-emergence of test positivity and symptoms after completion of oral outpatient SARS-CoV-2 treatments(35). This may have societal impacts related to extension of isolation along with health effects and reduced quality of life with prolonged viral infection. The exact mechanisms of COVID-19 rebound are unknown, but it may reflect incomplete viral eradication at the completion of oral treatment. SARD patients have altered underlying immunity and are immunosuppressed, with some known to have prolonged viral shedding(22), so it is possible that immunosuppressed patients could have higher risk of COVID-19 rebound. We found that 8% of SARD patients that received oral outpatient treatments experienced documented COVID-19 rebound. Since our study was retrospective, requiring documentation of recurrent positive COVID-19 test results and symptoms to confirm rebound cases, 8% of COVID-19 rebound among SARDs is likely an underestimate. Notably, no SARD patients in our study that experienced documented COVID-19 rebound were subsequently hospitalized, which is reassuring. Overall, this highlights the need for further research on COVID-19 in this vulnerable population, including prospective ascertainment of COVID-19 rebound, possible relationships with severe COVID-19(36) and long-COVID(37), and consideration for longer courses of oral regimens.

In addition to nirmatrelvir/ritonavir, monoclonal antibodies have been frequently used to treat COVID-19. They were the first outpatient treatment option shown to be effective in preventing severe COVID-19 among high-risk patients(1). Pre-exposure prophylaxis with monoclonal antibodies may also reduce severe outcomes, and among SARD patients who received B cell depleting therapy, monoclonal antibodies may be effective, even after vaccination(31, 38). Thus, our study adds to the literature by investigating all SARD patients, not only those at highest risk due to B cell depletion with resultant impaired humoral immunity. Compared to no outpatient treatment, monoclonal antibodies were associated with 80% lower odds of severe COVID-19. Since patients that received monoclonal antibodies may have had high clinical suspicion to progress to severe COVID-19, this is unlikely to explain lower odds of severe COVID-19. Indeed, SARD patients who received monoclonal antibodies had more comorbidities and worse kidney function than those without treatment. Even with oral options available, many clinicians and SARD patients may choose to receive monoclonal antibodies, due to contraindications for use of nirmatrelvir/ritonavir or to avoid COVID-19 rebound after oral medications. In the analysis that compared nirmatrelvir/ritonavir and monoclonal antibodies, there was no statistical difference, suggesting both may be similarly effective.

Strengths of our study include the contemporary nature of the cohort that included recently-approved oral outpatient SARS-CoV-2 treatment options and the systematic approach to COVID-19 case ascertainment in patients with SARDs. The algorithm we used to identify SARD patients has high validity and identified patients on immunosuppression, those at highest risk of severe COVID-19 outcomes. We also performed medical record review that confirmed outpatient SARS-CoV-2 treatment status and COVID-19 rebound, previously not studied systematically among SARDs. We were able to measure important factors related to treatment use and outcomes, including comorbidities, kidney function, immunomodulating medications, vaccine status, and tixagevimab/cilgavimab use.

There are limitations to consider. First, the analysis was performed using data from a single geographic area with a high vaccination rate so it may not generalize to other settings. However, we still detected statistical differences in severe COVID-19 risk that may be even more pronounced in less vaccinated populations. Second, we may not have identified some people with COVID-19 who diagnosed themselves at home with rapid antigen tests. However, higher risk people may be more likely to seek testing and treatment, and this may have therefore biased our findings toward the null. Third, some of the severe COVID-19 outcomes may have been due to incidentally-diagnosed COVID-19 from screening during hospitalizations for other reasons or could have been nosocomial infections. Our findings remained robust in a sensitivity analysis that required a time separation between the index date and outcome. Fourth, it is possible that the results may have been affected by unmeasured confounding that may include specific contraindications to nirmatrelvir/ritonavir, social determinants of health, and access to care. Finally, we relied on medical documentation to identify cases of COVID-19 rebound. It is possible that some patients experienced this, and it was not documented. Therefore, we presented only descriptive studies, and this estimate should be viewed as conservative.

In conclusion, we found that outpatient SARS-CoV-2 treatment was associated with strongly reduced odds of severe COVID-19 compared to no treatment. Over time, more SARD patients were treated for COVID-19 as outpatients, mostly with nirmatrelvir/ritonavir or monoclonal antibodies. The proportion of SARDs experiencing confirmed COVID-19 rebound was at least 8%, a conservative estimate due to the stringent definition we used that required documentation. These findings should encourage outpatient SARS-CoV-2 treatment among SARD patients.

## Data Availability

Data are available upon reasonable request and with appropriate institutional review board approval.

## Acknowledgements

None.

## Contributors

GQ, ZSW, and JAS designed the study, were responsible for the acquisition, analysis, and interpretation of the data, and drafted and revised the article. XW was involved in the analysis and interpretation of the data. GQ, XW, NJP, YK, XF, CEC, KMMV, ENK, EPB, KJB, SS, ZKW, DJT, MEW, ZSW, JAS were involved in the data acquisition, interpretation, and revision of the manuscript. ZSW and JAS are joint senior authors. All authors approved the final version of the article. JAS accepts full responsibility for the work and the conduct of the study, had access to the data, and controlled the decision to publish.

## Ethics approval

This study was approved by the Mass General Brigham Institutional Review Board (2020P000840).

## Patient and public involvement

Patients were not involved in the design, conduct or reporting of this study.

