## Supplemental Table 1 for "Outcomes with and without outpatient SARS-CoV-2 treatment for patients with COVID-19 and systemic autoimmune rheumatic diseases: A retrospective cohort study"

**Supplementary Table S1**. Rheumatic disease terms used to identify patients with systemic rheumatic disease and their associated ICD-10 codes.

| **Category** | **Rheumatic Disease (ICD-10 codes)** |
| --- | --- |
| Inflammatory arthritis | - Rheumatoid arthritis (M05%, M06%) - Inflammatory arthritis or inflammatory polyarthropathy (M06.4) - Juvenile idiopathic arthritis (M08.20) - Psoriatic arthritis or arthropathic psoriasis (L40.50) - Ankylosing spondylitis (M45.9) |
| Vasculitis | - Anti-neutrophil cytoplasmic antibody-associated vasculitis: granulomatosis with polyangiitis, eosinophilic granulomatosis with polyangiitis, microscopic polyangiitis (M31.3, M31.7, M30.0) - Kawasaki disease (M30.3) - Takayasu arteritis (M31.4) - Polyarteritis nodosa (M30.0) - Giant cell arteritis (M31.6) - Polymyalgia rheumatica (M35.3) - Behçet disease (M35.2) - Unspecified arteritis (I77.6) |
| Other Systemic Autoimmune Diseases | - Systemic lupus erythematosus (M32%) - Sjogren’s syndrome (M35.0) - Idiopathic inflammatory myositis: dermatomyositis, polymyositis, statin-associated autoimmune myositis, unspecified myositis (G72.49, G72.41, M33) - Systemic sclerosis (M34.0, M34.1, M34.8%, M34.9) - Mixed connective tissue disease (M35.1) - Antiphospholipid syndrome (D68.61) |

ICD = international classification of disease, 10^th^ revision codes. % indicates a wild character to capture all instances of a given code.

**Supplementary Table S2**. Immunomodulatory medications used to identify patients with systemic rheumatic disease

| **Category and generic name** | **Brand name(s)** | **Mechanism of action (for targeted therapy)** | **Route** | **CPT code if intravenous** |
| --- | --- | --- | --- | --- |
| **Glucocorticoids** |  |  |  |  |
| Prednisone (minimum of 30 pills) | Deltasone  Prednicot  Prednisone Intensol  Rayos  Sterapred  Sterapred DS |  | Oral |  |
| Methylprednisolone (minimum of 30 pills) | Medrol |  | Oral |  |
| **Conventional Synthetic DMARDs** |  |  |  |  |
| Azathioprine | Imuran  Azasan |  | Oral |  |
| Methotrexate | Otrexup  Rasuvo  Rheumatrex  Trexall |  | Oral or subQ |  |
| Leflunomide | Arava |  | Oral |  |
| Mycophenolic acid | CellCept |  | Oral |  |
| Mycophenolate mofetil | Myfortic |  | Oral |  |
| Sulfasalazine | Azulfidine |  | Oral |  |
| Hydroxychloroquine | Plaquenil |  | Oral |  |
| Chloroquine | Aralen |  | Oral |  |
| **Targeted Synthetic DMARDs** |  |  |  |  |
| Tofacitinib | Xeljanz | JAK inhibitor | Oral |  |
| Baricitinib | Olumiant | JAK inhibitor | Oral |  |
| Upadacitinib | Rinvoq | JAK inhibitor | Oral |  |
| **Biologic DMARDs** |  |  |  |  |
| Rituximab | Rituxan  Truxima  Ruxience | Anti-CD20 monoclonal antibody | IV | J9310  Q5115  Q5119 |
| Ocrelizumab | Ocrevus | Anti-CD20 monoclonal antibody | IV | J2350 |
| Abatacept | Orencia | CTLA-4 Ig | subQ or IV | J0129 |
| Infliximab | Remicade  Inflectra  Renflexis  Avsola | TNF inhibitor | IV | J1745  Q5103 Q5104  Q5121 |
| Etanercept | Enbrel | TNF inhibitor | subQ |  |
| Adalimumab | Humira | TNF inhibitor | subQ |  |
| Certolizumab | Cimzia | TNF inhibitor | subQ or IV | J0717 |
| Golimumab | Simponi | TNF inhibitor | subQ or IV | J1602 |
| Anakinra | Kineret | IL-1 inhibitor | subQ or IV | n/a (coded under “other biologic” J3490 or J3590 but doesn’t seem to have its own) |
| Canakinumab | Ilaris | IL-1 inhibitor | subQ or IV | J0638 |
| Mepolizumab | Nucala | IL-5 inhibitor | subQ or IV | J2182 |
| Benralizumab | Fasenra | IL-5 inhibitor | subQ or IV | J0517 |
| Tocilizumab | Actemra | IL-6 inhibitor | subQ or IV | J3262 |
| Sarilumab | Kevzara | IL-6 inhibitor | subQ |  |
| Secukinumab | Cosentyx | IL-17A inhibitor | subQ |  |
| Ixekizumab | Taltz | IL-17A inhibitor | subQ |  |
| Ustekinumab | Stelara | IL-12/23 inhibitor | subQ or IV | J3358 |
| Guselkumab | Tremfya | IL-23 inhibitor | subQ |  |
| Belimumab | Benlysta | BLyS inhibitor | subQ or IV | J0490 |
| Eculizumab | Soliris | C5 inhibitor | IV | J1300 |
| **Other** |  |  |  |  |
| Tacrolimus | Prograf  Envarsus  Astagraf  Hecoria |  | Oral or IV | J7525 |
| Cyclosporine | Gengraf  Neoral  Sandimmune |  | Oral or IV | J7516 |
| Apremilast | Otezla | PDE4 inhibitor | Oral |  |
| Cyclophosphamide | Cytoxan |  | Oral or IV | J9070 |

CPT, Current Procedural Terminology; CTLA-4, cytotoxic T-lymphocyte-associated protein 4; DMARD, disease-modifying anti-rheumatic drug; IL, interleukin; JAK, Janus kinase; PDE, phosphodiesterase; TNF, tumor necrosis factor

**Supplemental Table 3**. Baseline characteristics for SARD patients at COVID-19 onset among molnupiravir, outpatient remdesivir, or combination users.

| **Characteristic** | **Molnupiravir use (n=5)** | **Outpatient remdesivir use (n=3)** | **Combination use (n=6)** |
| --- | --- | --- | --- |
| **Demographics** |  |  |  |
| Age (mean, SD, years) | 68.26 (9.7) | 65.78 (17.4) | 55.78 (6.8) |
| Female | 4 (80.0%) | 3 (100.0%) | 6 (100.0%) |
| Race |  |  |  |
| White | 5 (100.0%) | 3 (100.0%) | 5 (83.3%) |
| Black or African American | 0 (0.0%) | 0 (0.0%) | 0 (0.0%) |
| Asian | 0 (0.0%) | 0 (0.0%) | 1 (16.7%) |
| Hispanic or Latinx ethnicity | 0 (0.0%) | 0 (0.0%) | 0 (0.0%) |
| **Lifestyle** |  |  |  |
| Body mass index (mean, SD, kg/m^2^) | 30.00 (6.2) | 30.14 (7.2) | 33.51 (13.8) |
| Smoking status |  |  |  |
| Never | 2 (40.0%) | 2 (66.7%) | 4 (66.7%) |
| Past | 3 (60.0%) | 1 (33.3%) | 2 (33.3%) |
| Current | 0 (0.0%) | 0 (0.0%) | 0 (0.0%) |
| **Comorbidities** |  |  |  |
| Charlson Comorbidity Index categories |  |  |  |
| 0 | 1 (20.0%) | 0 (0.0%) | 0 (0.0%) |
| 1 | 1 (20.0%) | 1 (33.3%) | 2 (33.3%) |
| 2 | 1 (20.0%) | 0 (0.0%) | 1 (16.7%) |
| ≥3 | 2 (40.0%) | 2 (66.7%) | 3 (50.0%) |
| Individual comorbidities |  |  |  |
| Hypertension | 3 (60.0%) | 2 (66.7%) | 2 (33.3%) |
| Diabetes | 1 (20.0%) | 2 (66.7%) | 2 (33.3%) |
| Coronary artery disease | 2 (40.0%) | 0 (0.0%) | 2 (33.3%) |
| Heart failure | 2 (40.0%) | 1 (33.3%) | 0 (0.0%) |
| Asthma | 0 (0.0%) | 1 (33.3%) | 2 (33.3%) |
| Chronic obstructive pulmonary disease | 1 (20.0%) | 0 (0.0%) | 0 (0.0%) |
| Obstructive sleep apnea | 0 (0.0%) | 2 (66.7%) | 2 (33.3%) |
| Chronic kidney disease | 0 (0.0%) | 1 (33.3%) | 2 (33.3%) |
| Malignancy excluding non-melanoma skin cancer | 1 (20.0%) | 0 (0.0%) | 1 (16.7%) |
| Interstitial lung disease | 0 (0.0%) | 0 (0.0%) | 0 (0.0%) |
| **Rheumatic disease diagnosis** |  |  |  |
| Rheumatoid arthritis | 1 (20.0%) | 0 (0.0%) | 3 (50.0%) |
| Psoriatic arthritis | 1 (20.0%) | 0 (0.0%) | 0 (0.0%) |
| Giant cell arteritis and/or polymyalgia rheumatica | 0 (0.0%) | 1 (33.3%) | 0 (0.0%) |
| Systemic lupus erythematosus | 1 (20.0%) | 1 (33.3%) | 2 (33.3%) |
| ANCA-associated vasculitis and other miscellaneous vasculitis | 0 (0.0%) | 0 (0.0%) | 1 (16.7%) |
| Axial spondyloarthritis | 1 (20.0%) | 0 (0.0%) | 0 (0.0%) |
| Mixed connective tissue disease | 1 (20.0%) | 0 (0.0%) | 0 (0.0%) |
| **Immunomodulatory medications** |  |  |  |
| Oral glucocorticoid | 0 (0.0%) | 0 (0.0%) | 0 (0.0%) |
| Biologic DMARDs |  |  |  |
| Anti-CD20 monoclonal antibody | 0 (0.0%) | 0 (0.0%) | 3 (50.0%) |
| TNF inhibitor | 3 (60.0%) | 0 (0.0%) | 0 (0.0%) |
| IL-6 receptor inhibitor | 0 (0.0%) | 1 (33.3%) | 1 (16.7%) |
| Conventional synthetic DMARDs |  |  |  |
| Hydroxychloroquine | 2 (40.0%) | 2 (66.7%) | 3 (50.0 %) |
| Mycophenolate mofetil/mycophenolic acid | 0 (0.0%) | 0 (0.0%) | 1 (16.7%) |
| Leflunomide | 0 (0.0%) | 0 (0.0%) | 1 (16.7%) |
| Azathioprine | 0 (0.0%) | 0 (0.0%) | 1 (16.7%) |
| Cyclosporine | 1 (20.0%) | 0 (0.0%) | 0 (0.0%) |
| Tacrolimus | 0 (0.0%) | 0 (0.0%) | 1 (16.7%) |
| **Previous COVID-19 immunity** |  |  |  |
| Vaccination status |  |  |  |
| Unvaccinated | 0 (0.0%) | 0 (0.0%) | 0 (0.0%) |
| Partially vaccinated | 0 (0.0%) | 0 (0.0%) | 0 (0.0%) |
| 2 doses mRNA or 1 dose J&J | 1 (20.0%) | 0 (0.0%) | 1 (16.7%) |
| Additional doses | 4 (80.0%) | 3 (100.0%) | 5 (83.3%) |
| Tixagevimab/cilgavimab use | 0 (0.0%) | 0 (0.0%) | 1 (16.7%) |
| Previous COVID-19 infection | 0 (0.0%) | 0 (0.0%) | 0 (0.0%) |

COVID-19, coronavirus disease 2019; eGFR, estimated glomerular filtration rate; IQR, interquartile range; SARD, systemic autoimmune rhematic disease; SD, standard deviation.

**Supplemental Table 4**. Severe COVID-19 outcomes by outpatient treatment among SARD patients (n=704), only considering outcomes that occur at least one day after initial positive SARS-CoV-2 test.

| **Outcome** | **All Rheumatic Disease Patients with COVID-19 (n=704)** | **No outpatient treatment (n=278)** | **Any treatment**  **(n=426)** | **Nirmatrelvir/**  **ritonavir use (n=307)** | **Monoclonal antibody use (n=105)** |
| --- | --- | --- | --- | --- | --- |
| Hospitalization | 38 (5.4%) | 29 (10.4%) | 9 (2.1%) | 4 (1.3%) | 5 (4.8%) |
| Death | 3 (0.4%) | 2 (0.7%) | 1 (0.2%) | 1 (0.3%) | 0 (0.0%) |
| Severe COVID-19 (hospitalization or death) | 39 (5.5%) | 30 (10.8%) | 9 (2.1%) | 4 (1.3%) | 5 (4.8%) |

*There were no severe COVID-19 outcomes among molnupiravir (n=5), remdesivir (n=3), or combination (n=6; 4 received nirmatrelvir/ritonavir and monoclonal antibodies and 2 received molnupiravir and monoclonal antibodies) users.

COVID-19, coronavirus disease 2019; SARD, systemic autoimmune rheumatic disease; SARS-CoV-2, severe acute respiratory syndrome coronavirus 2.

**Supplemental Table 5**. Odds ratios for severe COVID-19 (hospitalization or death) by outpatient treatment status, only considering outcomes that occur at least one day after initial positive SARS-CoV-2 test.

| **Comparisons (reference=second group listed)** | **Unadjusted OR for severe COVID-19 (95%CI)** | **Multivariable* OR for severe COVID-19 (95%CI)** |
| --- | --- | --- |
| **Primary analysis** |  |  |
| Any outpatient treatment vs. no outpatient treatment | 0.18 (0.08, 0.38) | 0.22 (0.10, 0.48) |
| **Secondary analyses** |  |  |
| Nirmatrelvir/ritonavir vs. no outpatient treatment | 0.11 (0.04, 0.31) | 0.15 (0.05, 0.43) |
| Monoclonal antibodies vs. no outpatient treatment | 0.41 (0.16, 1.10) | 0.41 (0.15, 1.11) |
| Nirmatrelvir/ritonavir vs. all others | 0.14 (0.05, 0.39) | 0.19 (0.06, 0.54) |
| Monoclonal antibodies vs. all others | 0.83 (0.32, 2.18) | 0.67 (0.24, 1.83) |
| Nirmatrelvir/ritonavir vs. monoclonal antibodies | 0.26 (0.07, 1.00) | 0.46 (0.11, 1.97) |

*Adjusted for continuous age, continuous Charlson Comorbidity Index, continuous estimated glomerular filtration rate, and race.

CI, confidence interval; COVID-19, coronavirus disease 2019; OR, odds ratio; SARS-CoV-2, severe acute respiratory syndrome coronavirus 2.

**Supplemental Table 6**. Baseline characteristics stratified by COVID-19 rebound among users of nirmatrelvir/ritonavir (n=311).

| **Characteristic** | **Nirmatrelvir/**  **ritonavir use and documented rebound (n=24)** | **Nirmatrelvir/**  **ritonavir use and no documented rebound (n=287)** |
| --- | --- | --- |
| **Demographics** |  |  |
| Mean age (SD), years | 54.29 (13.4) | 57.32 (15.0) |
| Female | 20 (83.3%) | 219 (76.3%) |
| Race |  |  |
| White | 20 (83.3%) | 242 (84.3%) |
| Black or African American | 0 (0.0%) | 15 (5.2%) |
| Asian | 2 (8.3%) | 8 (2.8%) |
| Other (Two or more) | 2 (8.3%) | 14 (4.9%) |
| Unknown | 0 (0.0%) | 8 (2.8%) |
| Hispanic or Latinx ethnicity | 0 (0.0%) | 4 (1.4%) |
| **Lifestyle** |  |  |
| Body mass index (mean, SD, kg/m^2^) | 24.72 (7.3) | 28.13 (7.4) |
| Categorical BMI |  |  |
| <18.5 (underweight) | 0 (0.0%) | 9 (3.1%) |
| 18.5 to <25 (normal) | 11 (45.8%) | 94 (32.8%) |
| 25 to <30 (overweight) | 8 (33.3%) | 84 (29.3%) |
| ≥30 (obese) | 4 (16.7%) | 97 (33.8%) |
| Missing | 1 (4.2%) | 3 (1.1%) |
| Smoking status |  |  |
| Never | 16 (66.7%) | 189 (65.9%) |
| Past | 7 (29.2%) | 86 (30.0%) |
| Current | 0 (0.0%) | 9 (3.1%) |
| Unknown | 1 (4.2%) | 3 (1.1%) |
| **Comorbidities** |  |  |
| Median Charlson Comorbidity Index (IQR) | 1 (1, 3) | 1 (1, 2) |
| 0 | 4 (16.7%) | 43 (15.0%) |
| 1 | 9 (37.5%) | 144 (50.2%) |
| 2 | 4 (16.7%) | 47 (16.4%) |
| ≥3 | 7 (29.2%) | 53 (18.5%) |
| Individual comorbidities |  |  |
| Hypertension | 4 (16.7%) | 99 (34.5%) |
| Diabetes | 3 (12.5%) | 17 (5.9%) |
| Coronary artery disease | 2 (8.3%) | 22 (7.7%) |
| Heart failure | 0 (0.0%) | 4 (1.4%) |
| Asthma | 5 (20.8%) | 39 (13.6%) |
| Chronic obstructive pulmonary disease | 0 (0.0%) | 4 (1.4%) |
| Obstructive sleep apnea | 2 (8.3%) | 26 (9.1%) |
| Chronic kidney disease | 5 (20.8%) | 23 (8.0%) |
| Interstitial lung disease | 0 (0.0%) | 9 (3.1%) |
| Malignancy excluding non-melanoma skin cancer | 1 (4.2%) | 34 (11.9%) |
| Non-melanoma skin cancer | 0 (0.0%) | 9 (3.1%) |
| **SARD diagnosis** |  |  |
| Rheumatoid arthritis | 10 (41.7%) | 149 (51.9%) |
| Psoriatic arthritis | 5 (20.8%) | 57 (19.9%) |
| Giant cell arteritis and/or polymyalgia rheumatic | 0 (0.0%) | 13 (4.5%) |
| Systemic lupus erythematosus | 5 (20.8%) | 33 (11.5%) |
| Sjogren’s syndrome | 2 (8.3%) | 5 (1.7%) |
| ANCA-associated vasculitis and other miscellaneous vasculitis | 0 (0.0%) | 9 (3.1%) |
| Systemic sclerosis | 0 (0.0%) | 4 (1.4%) |
| Axial spondyloarthritis | 1 (4.2%) | 4 (1.4%) |
| Mixed connective tissue disease | 0 (0.0%) | 2 (0.7%) |
| Antiphospholipid antibody syndrome | 0 (0.0%) | 3 (1.1%) |
| Behcet disease | 1 (4.2%) | 2 (0.7%) |
| Idiopathic inflammatory myositis | 0 (0.0%) | 0 (0.0%) |
| Takayasu arteritis | 0 (0.0%) | 1 (0.4%) |
| Juvenile idiopathic arthritis | 0 (0.0%) | 1 (0.4%) |
| Multiple primary rheumatic diseases | 0 (0.0%) | 4 (1.4%) |
| **Immunomodulatory medications** |  |  |
| Oral glucocorticoid | 0 (0.0%) | 6 (2.1%) |
| Conventional synthetic DMARDs | 18 (75.0%) | 190 (66.2%) |
| Hydroxychloroquine | 11 (45.8%) | 76 (26.5%) |
| Hydroxychloroquine monotherapy | 6 (25.0%) | 37 (12.9%) |
| Methotrexate | 4 (16.7%) | 102 (35.5%) |
| Mycophenolate mofetil/mycophenolic acid | 0 (0.0%) | 14 (4.9%) |
| Leflunomide | 2 (8.3%) | 22 (7.7%) |
| Azathioprine | 3 (12.5%) | 6 (2.1%) |
| Sulfasalazine | 0 (0.0%) | 15 (5.2%) |
| Apremilast | 0 (0.0%) | 2 (0.7%) |
| Cyclophosphamide | 0 (0.0%) | 4 (1.4%) |
| Cyclosporine | 0 (0.0%) | 6 (2.1%) |
| Tacrolimus | 2 (8.3%) | 8 (2.8%) |
| Biologic DMARDs | 8 (33.3%) | 128 (44.6%) |
| Anti-CD20 monoclonal antibody | 0 (0.0%) | 15 (5.2%) |
| TNF inhibitor | 6 (25.0%) | 80 (27.9%) |
| IL-6 receptor inhibitor | 0 (0.0%) | 11 (3.8%) |
| B-cell activating factor inhibitor | 0 (0.0%) | 1 (0.4%) |
| IL-23 inhibitor | 0 (0.0%) | 1 (0.4%) |
| IL-17 inhibitor | 2 (8.3%) | 11 (3.8%) |
| IL-12/IL-23 inhibitor | 0 (0.0%) | 1 (0.4%) |
| IL-1 inhibitor | 0 (0.0%) | 1 (0.4%) |
| CTLA-4 immunoglobulin | 0 (0.0%) | 8 (2.8%) |
| Targeted synthetic DMARD |  |  |
| JAK inhibitor | 0 (0.0%) | 8 (2.8%) |
| **Previous COVID-19 immunity** |  |  |
| Vaccination status |  |  |
| Unvaccinated | 0 (0.0%) | 8 (2.8%) |
| Partially vaccinated | 0 (0.0%) | 0 (0.0%) |
| 2 doses mRNA or 1 dose J&J | 3 (12.5%) | 38 (13.2%) |
| Additional doses | 21 (87.5%) | 241 (84.0%) |
| Tixagevimab/cilgavimab use | 0 (0.0%) | 6 (2.1%) |
| Previous COVID-19 infection | 0 (0.0%) | 8 (2.8%) |

ANCA, antineutrophil cytoplasmic antibodies; COVID-19, coronavirus disease 2019; CTLA-4, cytotoxic T-lymphocyte-associated protein 4; DMARDs, disease-modifying antirheumatic drugs; eGFR, estimated glomerular filtration rate; IL, interleukin; IQR, interquartile range; SARD, systemic autoimmune rheumatic disease; SD, standard deviation; TNF, tumor necrosis factor.
